# Automated and Interpretable Detection of Hippocampal Sclerosis in temporal lobe epilepsy: AID-HS

**DOI:** 10.1101/2023.10.13.23296991

**Authors:** Mathilde Ripart, Jordan DeKraker, Maria H Eriksson, Rory J. Piper, Jia-Jie Mo, Ting-Yu Su, Ryuzaburo Kochi, Irene Wang, Gavin P Winston, Chris A Clark, Felice D’Arco, Kshitij Mankad, Ali R. Khan, Torsten Baldeweg, Sophie Adler, Konrad Wagstyl

**Affiliations:** UCL Great Ormond Street Institute of Child Health, London, UK; McGill University, Montreal, Canada; Great Ormond Street Hospital, London; Beijing Tiantan Hospital, Beijing, China; Epilepsy Center, Neurological Institute, Cleveland Clinic, Cleveland, USA; Queen’s University, Kingston, Canada; UCL Queen Square Institute of Neurology, London, UK; Department of Medical Biophysics, Schulich School of Medicine and Dentistry, Canada; Wellcome Centre for Human Neuroimaging, University College London, London, UK

**Keywords:** epilepsy, hippocampal sclerosis, machine learning, structural MRI

## Abstract

Hippocampal Sclerosis (HS) can elude visual detection on MRI scans of patients with temporal lobe epilepsy (TLE), causing delays in surgical treatment and reducing the likelihood of postsurgical seizure-freedom. We developed an open-source software that (1) detects HS from structural MRI scans, (2) generalises across a heterogeneous multicentre cohort of children and adults, and (3) generates individualised predictions for clinical evaluation.

We included a cohort of 363 participants (152 patients with HS, 90 disease controls with focal cortical dysplasia, and 121 healthy controls) from four epilepsy centres in the UK, North America, and China. We used the open-source software HippUnfold to extract morphological surface-based features and volumes of the hippocampus from T1w MRI scans. We compared pathological hippocampal morphology in patients with HS to normative growth charts generated from healthy controls, and to the contralateral hippocampi in patients with HS. HS was characterised by decreased volume, thickness and gyrification as well as increased mean and intrinsic curvature. A logistic regression classifier trained on these features detected 90.1% of HS patients, and accurately lateralised 97.4% of the HS cohort. Crucially, in patients with MRI-negative histopathologically confirmed HS, the classifier detected HS in 79.2% (19/24) and accurately lateralised the lesions in 91.7% (22/24). The Automated and Interpretable Detection of Hippocampal Sclerosis classifier (AID-HS) was packaged into an open-source pipeline, which detects and lateralises HS and generates individualised patient reports that characterise hippocampal developmental abnormalities.

AID-HS is capable of accurately detecting and lateralising HS in a large, heterogeneous, multi-centre, cohort of paediatric and adult patients with diagnostically challenging HS. Moreover, by offering transparent, robust and interpretable results, AID-HS can support the presurgical evaluation of patients with suspected TLE.

## Introduction

Hippocampal sclerosis (HS) is the leading cause of refractory focal epilepsy in adults, and the third most common cause in children^1,2^. It is amenable to surgical resection, with seizure freedom reported in 76% of individuals at one year after surgery and 70% after five years^3^. HS is typically diagnosed using structural MRI and is characterised by atrophy (i.e., volume reduction) of the affected hippocampus on T1-weighted MRI scans, alongside hippocampal T2/FLAIR hyperintensity^4,5^. Additionally, there is evidence of shape abnormalities in the affected hippocampus, such as increased curvature of the tail^6^ and a reduction in hippocampal dentations^7^.

Despite its characteristic features, MRI abnormalities can be subtle, and HS is reported to represent approximately 10% of focal epilepsy cases that escape detection during routine visual inspection of MRI scans^8,9^. Critically, individuals with an “MRI-negative” scan have significantly lower rates of postsurgical seizure freedom (45%) compared to those with identified lesions (72%-81%)^10^. Attempts to localise MRI abnormalities in these patients often involve additional investigations, such as Positron Emission Tomography (PET) imaging or invasive intracranial EEG. Alongside inducing delays to surgical resection, these procedures place an additional burden on patients and families^11,12^. Developing imaging software that can help to detect and characterise subtle cases of HS on presurgical MRI scans has the potential to streamline the surgical treatment pathway for these patients and improve their postsurgical outcomes.

Machine-learning technology is increasingly used to improve the detection of epilepsy-associated abnormalities on MRI, including HS^13^. Past studies have trained models on volumetric- or surface-based features of the hippocampus and the adjacent temporal neocortex to both distinguish patients with HS from healthy controls (i.e., detection) and lateralise the side of HS-associated abnormalities (i.e., lateralisation)^14–18^. However, these models have often been trained on small, single-centre datasets^14–16^ and are therefore unlikely to generalise well to other centres with different patient cohorts, MRI hardware and scanning protocols. Moreover, models trained on larger multi-centre datasets have been comprised of exclusively adult patients^17,18^, thus limiting their applicability in paediatric patients. Finally, and perhaps most importantly, the models published to date have lacked open-source code, and can therefore not be independently validated by other centres nor used for clinical evaluation.

The development of automated tools that can effectively accommodate for both adult and paediatric patients is challenging, primarily due to the ongoing maturation of the hippocampus during childhood – notably its increase in volume, infolding, asymmetries, and myelination^19–21^. Possible solutions to this challenge may involve measuring asymmetries within individuals, as this has shown to effectively correct for the effects of age and sex^22,23^. Another solution may involve the use of normative charts^24^. Normative charts for the hippocampus have previously been used to characterise typical and pathological morphological changes that occur with ageing^25^ and to detect pathological changes in Alzheimer’s disease^26^. Its use remains unexplored in HS, where it could help characterise hippocampal abnormalities across development.

We aimed to bridge the current gap by creating an open-source software for Automated and Interpretable Detection of Hippocampal Sclerosis in patients with epilepsy (AID-HS). This software leveraged a large, heterogenous cohort of adult and paediatric patients from four epilepsy centres across the UK, North America, and China to increase its applicability across diverse patients and clinical settings. To ensure interpretability of the results, we extracted comprehensive surface- and volumetric-based MRI features of the hippocampus using the open-source tool HippUnfold. We used these features to characterise HS abnormalities, through the analysis of normative trajectories and assessment of asymmetries, and to automate the detection and lateralisation of HS with machine-learning. Finally, AID-HS is shared as a user-friendly open-source software that automatically generates individualised and interpretable reports, to facilitate its clinical evaluation.

## Materials and methods

### Cohorts and MRI processing

#### Inclusion and exclusion criteria

Following local Institutional Review Board approval, data were retrospectively collected and anonymised from four epilepsy centres: Great Ormond Street Hospital (GOSH), UK; the National Hospital for Neurology and Neurosurgery (NHNN), UK; Beijing Tiantan Hospital (BTH), China; and Cleveland Clinic (CC), USA; prior to sharing with University College London. Patients were included if they had histopathologically confirmed HS. “MRI-negative” patients were lateralised following additional investigations, such as PET, single-photon emission computerized tomography (SPECT), video-telemetry or intracranial EEG. A cohort of patients with focal cortical dysplasia (FCD) were included as disease controls, alongside a healthy control cohort who had been scanned for research purposes. Patients and controls were included if they had a preoperative 3D T1w MRI scan acquired at 3T (Figure 1A) and were more than three years old at the time of MRI acquisition.

**Figure 1:**
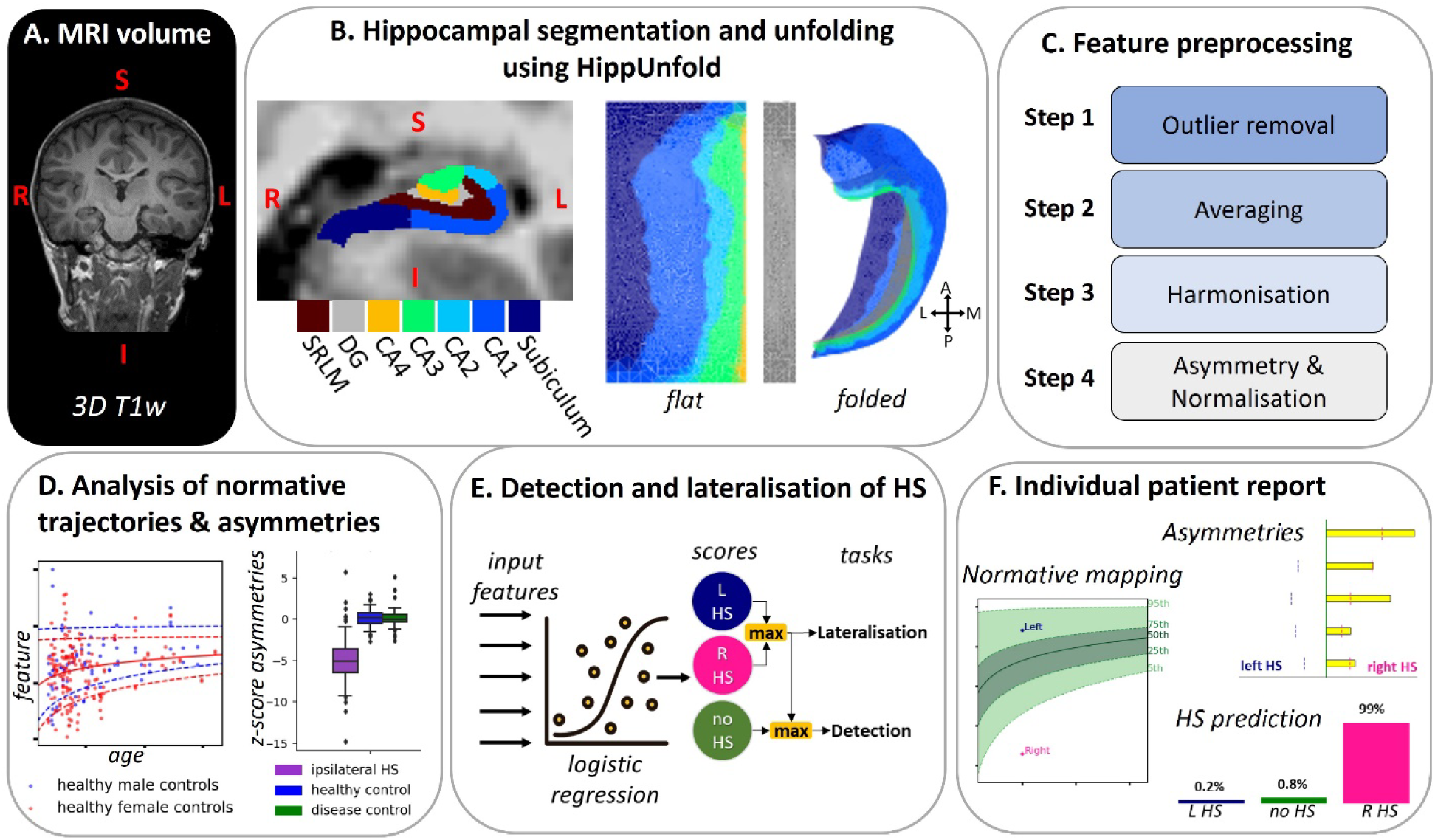
AID-HS overview. The T1w scan (A) is used as input in HippUnfold, which generates hippocampal segmentations and mesh surfaces that can be visualised flat or folded (B). Surface-based features undergo preprocessing to remove outliers, adjust for site-based batch effects, and account for inter and intra-individual differences (C). Affected hippocampal features are compared to i) normative developmental trajectories of hippocampal features generated from the healthy controls, and ii) contralateral hippocampi to characterise the asymmetries (D). Asymmetries are used to train a logistic regression model to predict the likelihood of an individual having a left HS, right HS or no HS. These scores are used to detect and lateralise HS (E). AID-HS outputs individualised patient reports that detail HS detection and lateralisation predictive scores as well as hippocampal feature asymmetries and characterisations of hippocampal abnormalities against normative trajectories (F). (SRLM: stratum radiatum, lacunosum, and moleculare)

#### Segmentation of the hippocampus

MRI data were manually quality controlled. Patients with large imaging artefacts (e.g., motion artefacts impairing the visibility of the anatomical structures) on their preoperative T1w MRI scan were excluded from further analysis.

T1w scans (Figure 1A) were used as input in the open-source software HippUnfold^27^ (Figure 1B). HippUnfold uses a U-Net neural network architecture to segment the hippocampal cortex, and applies Laplacian-based unfolding to fit hippocampal inner and outer hippocampal surface-meshes to the T1w images. Vertices in the meshes are assigned to the CA1-CA4 subfields, the subiculum, and the dentate gyrus (DG) using the BigBrain histological atlas^28^ in the unfolded space. HippUnfold also provides an automated quality check on the segmentation by examining dice overlap between the segmentation and a standard template. Following HippUnfold guidelines^27^, segmentations with a dice overlap score below 0.7 underwent visual inspection, and subjects with gross segmentation or surface errors were excluded.

Hippocampal volumes were also calculated using the open-source software FastSurfer^29^, a deep-learning accelerated version of FreeSurfer^30^, which uses a Convolutional Neural Network (CNN) architecture to segment cortical and subcortical structures including the hippocampus.

#### Extraction of hippocampal volume and surface-based features

We extracted the total volume (in mm^3^) from the hippocampi using both HippUnfold and FastSurfer hippocampal segmentations. For in-depth morphological analysis, we used three surface-based features – cortical thickness, gyrification, and curvature – extracted by HippUnfold at every vertex of the native folded hippocampal surface using Connectome Workbench (https://github.com/Washington-University/workbench). Cortical thickness was quantified as the distance between the white matter surface and the pial surface of the hippocampus. Curvature and gyrification measurements were obtained from a surface located at the mid-thickness between the inner and outer hippocampal surfaces. Mean and intrinsic (or Gaussian) curvatures were calculated for the outer surface of the hippocampus, and gyrification index was calculated as the ratio between the surface area in its native space and the unfolded space, which measures the degree of surface folding^31,32^. Finally, we extracted the total volume (in mm^3^) from the hippocampi using both HippUnfold and FastSurfer hippocampal segmentations.

#### Preprocessing of MRI features

Surface-based features underwent four steps of pre-processing (Figure 1C).

Step 1 Outlier removal: surface-based features underwent a filtering process to minimise the impact of isolated abnormal vertices. Vertices that fell outside five standard deviations of the mean distribution were replaced iteratively with the means of their neighbours. Subsequently, surface-based features were smoothed using a 1mm FWHM Gaussian kernel.

Step 2 Averaging: the mean of each surface-based feature was calculated for each hippocampus (including CA1-C4, subiculum, and DG), excluding 1% of vertices at both extremes of the anterior-posterior axis due to their significant variability^32^. This produced single mean values per hippocampus for each feature.

Step 3 Harmonisation: the averaged features were harmonised using neuroCombat^33^ to adjust for site-specific biases, without removing biological covariates (age, sex and disease status). The resulting features are henceforth referred to as “harmonised”.

Step 4 Asymmetry and Normalisation: asymmetry indexes, referred to as “asymmetries”, were computed to quantify differences between left (lh) and right (rh) hippocampi for all harmonised features following Equation 1. Each subject’s asymmetries were z-scored by the mean and standard deviation across healthy controls to account for typical asymmetry distributions following Equation 2. Output features of this process were referred to as “normalised”.

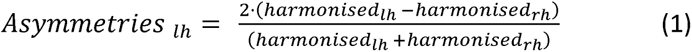

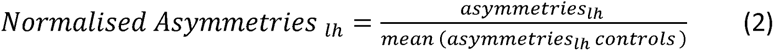

As hippocampal volumes from HippUnfold and FastSurfer were already extracted as averaged hippocampal values, only Steps 3-4 of the preprocessing were applied to them, after correcting them for intracranial volume (ICV) using a linear regression method^34^. A linear regression model was fit between FastSurfer-derived ICV and hippocampal volumes. The gradient of the regression model (*Grad*) and the mean intracranial volumes (*ICV_mean_*) of the healthy controls were used to compute the ICV-corrected hippocampal volumes (*Volume_hippo_*) for each participant (Equation 3).

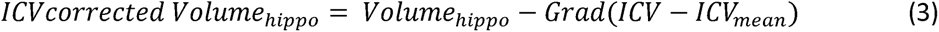

In the following methods and results “volume” will refer to the hippocampal volume derived from HippUnfold, if not otherwise specified.

### Healthy hippocampal anatomy

#### Evaluating HippUnfold in paediatric and adult data acquired at 3 Tesla

To evaluate the applicability of using HippUnfold to analyse paediatric and adult data acquired at 3T, we conducted a comparison between the surface-based features obtained in our cohort of healthy paediatric controls and adults and those previously computed from a cohort of young adults acquired at 7T from the Human Connectome Project (HCP) dataset, as detailed in DeKraker *et al*.^27^. To assess the vertex-level similarities between the two cohorts, Pearson’s correlation test was conducted, and correlations were corrected for multiple comparisons using the Holm method with a level of significance set at alpha=0.05.

#### Analysis of effect of age, sex and hemisphere on hippocampal features in controls

We used a linear regression model to quantify the effect of age, sex and hemisphere on the harmonised features in healthy controls (Table 1).

**Table 1:**
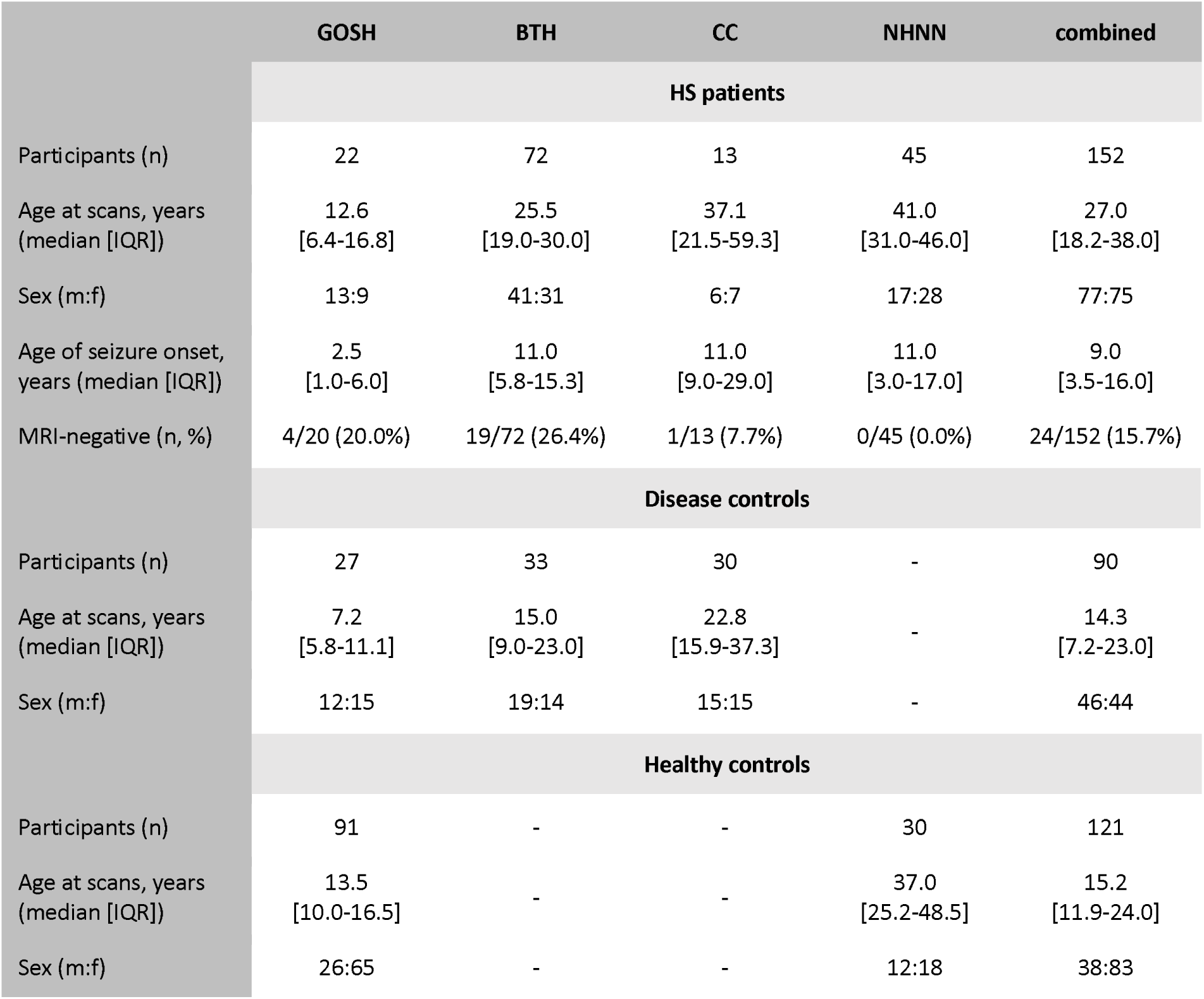
Demographic and clinical information for the cohort. (GOSH: Great Ormond Street Hospital, BTH: Beijing Tiantan Hospital, CC: Cleveland Clinic, NHNN: National Hospital for Neurology and Neurosurgery, IQR: interquartile range).

|  | GOSH | BTH | CC | NHNN | combined |
| --- | --- | --- | --- | --- | --- |
|  | **HS patients** |  |  |  |  |
| Participants (n) | 22 | 72 | 13 | 45 | 152 |
| Age at scans, years<br>(median [IQR]) | 12.6<br>[6.4-16.8] | 25.5<br>[19.0-30.0] | 37.1<br>[21.5-59.3] | 41.0<br>[31.0-46.0] | 27.0<br>[18.2-38.0] |
| Sex (m:f) | 13:9 | 41:31 | 6:7 | 17:28 | 77:75 |
| Age of seizure onset,<br>years (median [IQR]) | 2.5<br>[1.0-6.0] | 11.0<br>[5.8-15.3] | 11.0<br>[9.0-29.0] | 11.0<br>[3.0-17.0] | 9.0<br>[3.5-16.0] |
| MRI-negative (n, %) | 4/20 (20.0%) | 19/72 (26.4%) | 1/13 (7.7%) | 0/45 (0.0%) | 24/152 (15.7%) |
|  | **Disease controls** |  |  |  |  |
| Participants (n) | 27 | 33 | 30 | - | 90 |
| Age at scans, years<br>(median [IQR]) | 7.2<br>[5.8-11.1] | 15.0<br>[9.0-23.0] | 22.8<br>[15.9-37.3] | - | 14.3<br>[7.2-23.0] |
| Sex (m:f) | 12:15 | 19:14 | 15:15 | - | 46:44 |
|  | **Healthy controls** |  |  |  |  |
| Participants (n) | 91 | - | - | 30 | 121 |
| Age at scans, years<br>(median [IQR]) | 13.5<br>[10.0-16.5] | - | - | 37.0<br>[25.2-48.5] | 15.2<br>[11.9-24.0] |
| Sex (m:f) | 26:65 | - | - | 12:18 | 38:83 |

### Hippocampal anatomy in HS

#### Comparison of features in HS relative to normative growth charts

Normative growth curves for the healthy control harmonised features were generated using Generalized Additive Models (GAMs) accounting for age and sex. Estimates were visualised at the 5^th^, 50^th^ and 95^th^ percentiles of the healthy population. Additionally, percentile scores were calculated for the ipsilateral and contralateral hippocampi of each individual in the HS group, as well as for both hippocampi in the disease control group, and compared to the normative growth trajectories (Figure 1D).

#### Statistical analysis of hippocampal asymmetry

The distribution of the normalised asymmetries in the ipsilateral hippocampi of patients was compared with hippocampi from both disease controls and healthy controls, where the hemisphere in controls was randomly selected. Independent Welch t-tests (for normally distributed features) and Mann-Whitney tests (for non-normally distributed features) were conducted to compare the median normalised asymmetries among these three groups. P-values were corrected for multiple comparisons using the Holm method with a level of significance set at alpha=0.05.

A logistic regression was applied to each normalised asymmetries to find the threshold that best distinguish patients with right/left HS from healthy and disease controls. These abnormality thresholds were further used as benchmark for determining when normalised asymmetry is considered abnormal.

### Automated detection and lateralisation of HS

#### Classifier training and leave-one-site-out cross-validation

Features with significantly different normalised asymmetries in patients compared to controls were used to train a logistic regression classifier to detect and lateralise the side of the abnormality. The classifier was set up with a *multinomial* loss, the *lbfgs* solver, and balanced weights to account for multiclass and unbalanced labels. The classifier was trained to classify subjects into one of three classes – left HS, right HS, or no HS – and generated a score for each of these classes [S_LHS_, S_RHS,_ S_noHS_], which sum to 1. These scores were then used to assess the performances of the classifier at detecting (Equation 4) and lateralising the abnormalities (Equation 5). The classifier was evaluated using leave-one-site-out cross-validation, where a classifier was trained on the three sites withholding the final site for testing.

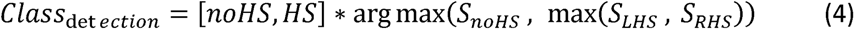

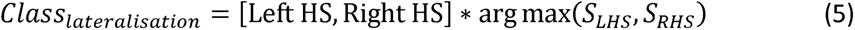

#### Classifier evaluation

Classifier performance was evaluated on two tasks: 1) detection – its ability to accurately distinguish patients with HS from healthy and disease controls 2) lateralisation – its ability to accurately lateralise the side of the abnormality in patients with HS (Figure 1E).

Additionally, classifier performance was stratified by age (children, adults), sex, histopathology (HS type-1, HS type-2, HS type-3, HS non-specified) and MRI status (MRI-negative, MRI-positive), and site in patients. Classifier performance was also compared between healthy and disease controls. These factors were tested as potential predictors of classifier accuracy using multivariable logistic regression models. Regression coefficients for each factor (β) and their significance (p-values) were reported alongside the performance breakdowns in Table 2.

**Table 2:** AID-HS classifier performance for the detection and lateralisation of HS. A breakdown of the performance is provided according to age, sex, MRI status, histopathology, and site. Multivariate logistic regression models were used to assess the impact of each factor (β coefficients) and the significance (*p-values*) on classifier performances at HS detection and lateralisation.

| | | N<br>subjects | Detection<br>(%) | Impact of factor<br>on detection<br>($\beta$ ; $p$-value) | Lateralisation<br>(%) | Impact of factor<br>on lateralisation<br>($\beta$ ; $p$-value) |
| --- | --- | --- | --- | --- | --- | --- |
| **Controls** |  | **211** | **94.3%** | - | - | - |
| group | Healthy controls | 121 | 95.9% | ref | - | - |
| | Disease controls | 90 | 92.2% | $\beta=-0.67$ ; $p=0.27$ | - | - |
| **HS patients** |  | **152** | **90.1%** | - | **97.4%** | - |
| Age | adult | 118 | 91.5% | ref | 98.3% | ref |
| | pediatric | 32 | 85.3% | $\beta=-0.36$ ; $p=0.68$ | 94.1% | $\beta=-0.11$ ; $p=0.94$ |
| Sex | male | 77 | 90.9% | ref | 97.4% | ref |
| | female | 75 | 89.3% | $\beta=-0.03$ ; $p=0.97$ | 97.3% | $\beta=0.16$ ; $p=0.89$ |
| MRI status | positive | 128 | 92.2% | ref | 98.4% | ref |
| | negative | 24 | 79.2% | $\beta=-1.59$ ; $p=0.07$ | 91.7% | $\beta=-1.37$ ; $p=0.28$ |
| Histopathology | HS type1 | 60 | 91.7% | ref | 100% | ref |
| | HS type 2 | 29 | 93.1% | $\beta=-0.13$ ; $p=0.90$ | 89.7% | $\beta=-30.4$ ; $p=1.0$ |
| | HS type 3 | 7 | 57.1% | $\beta=-1.52$ ; $p=0.14$ | 85.7 % | $\beta=-29.4$ ; $p=1.0$ |
| | non spec | 56 | 91.1% | $\beta=25.1$ ; $p=1.0$ | 100% | $\beta=-1.6$ ; $p=1.0$ |
| Site | BTH | 72 | 93.1% | ref | 95.8% | ref |
| | GOSH | 22 | 81.8% | $\beta=-1.28$ ; $p=0.22$ | 95.5% | $\beta=-1.36$ ; $p=0.50$ |
| | CC | 13 | 92.3% | $\beta=-1.60$ ; $p=0.21$ | 100% | $\beta=22.5$ ; $p=1.00$ |
| | NHNN | 45 | 88.9% | $\beta=-26.5$ ; $p=1.00$ | 100% | $\beta=-6.23$ ; $p=1.00$ |

#### Classifier comparison

Two further logistic regression classifiers with the same parametrisation were trained on hippocampal volumes extracted with FastSurfer. One classifier was trained on the volumes corrected for ICV, while the other was trained on the same volumes after applying Steps 3-4 of the pre-processing. This comparison allowed evaluation of the AID-HS classifier against a volumetric baseline, both with and without pre-processing techniques. Classifiers’ performances were compared in terms of accuracy at detecting and lateralising HS.

### Individual, interpretable hippocampal report

To provide an individualised and interpretable characterisation of hippocampal abnormalities, AID-HS generates individual reports for each subject (Figure 1F). The reports display HippUnfold segmentations and surface reconstructions, alongside automated quality control scores to highlight subjects in which the segmentation might have failed. Left and right hippocampal features are mapped against normative growth charts. Feature asymmetries are displayed to indicate the magnitude and direction of asymmetries, and compared to abnormality thresholds. Finally, the report includes the detection and lateralisation scores from the AID-HS classifier. The AID-HS reports have been co-designed with the neuroradiologists at GOSH to ensure they meet the clinical needs of providing transparent and comprehensive information that can aid in the diagnostic of patients with suspected HS.

## Data availability

All data analysis in this study was conducted using Python. The AID-HS software is openly available to download on GitHub (https://github.com/mathrip/AID-HS).

## Results

### Cohort

371 subjects were initially included in the study: 158 patients with HS, 91 disease controls with FCD, and 122 healthy controls. After quality control, one (0.3%) participant was excluded due to motion artefacts on their T1w scan, and seven (1.9%) participants failed the quality check on the HippUnfold segmentation.

The study’s final cohort consisted of 363 patients: 152 HS patients, 90 disease controls, and 121 healthy controls. The cohort included 200 adults (≥18 years old) and 163 children (<18 years old), with a median age of 27.0 years (IQR = 18.2-38.0) for patients, 14.3 years (IQR = 7.2-23.0) for disease controls, and 15.2 years (IQR = 11.9-24.0) for healthy controls (Table 1). The distribution of males and females was homogeneous in patients and disease controls (Table 1), while approximately two-thirds of the healthy controls were females (Chi-square (5, *N*=363) = 12.3, *p* = 0.03). In HS patients, the median age of epilepsy onset was 9.0 years old, and 15.7% of patients were categorised as MRI-negative. Among HS patients, the breakdown of histopathology diagnoses was 60 HS type-1, 29 HS type-2, 7 HS type-3 and 56 patients with non-specified HS.

### Healthy hippocampal anatomy

The surface-based thickness, gyrification and curvature features of the healthy controls exhibited a similar pattern to those obtained from the HCP cohort of young adults acquired at 7T (all *R*>0.89, all *p-values* < 0.001, Figure 2A), demonstrating the consistency of HippUnfold performance on 3T MRI. Developmental trajectories of harmonised features in healthy males and females are displayed in Figure 2B. Except for curvature, all features exhibited significant associations with age (all *p-values* < 0.05). Additionally, thickness and mean curvature were significantly different between males and females, and gyrification and intrinsic curvature were significantly different between the left and right hemispheres (all *p-values* <0.05) (Figure 2C). The linear regression gradient between the hippocampal volume and the ICV in healthy controls was 2e^-4^ (Figure 2D). Therefore, hippocampal volumes of each participant were corrected for ICV using Equation 3 with the parameters (*Grad*=2e^-4^, *ICV_mean_*=1.3e^6^mm^3^). Similarly, hippocampal volumes extracted from FastSurfer were corrected for ICV using the parameters (*Grad*=3e^-4^, *ICV_mean_*=1.3e^6^ mm^3^).

**Figure 2:**
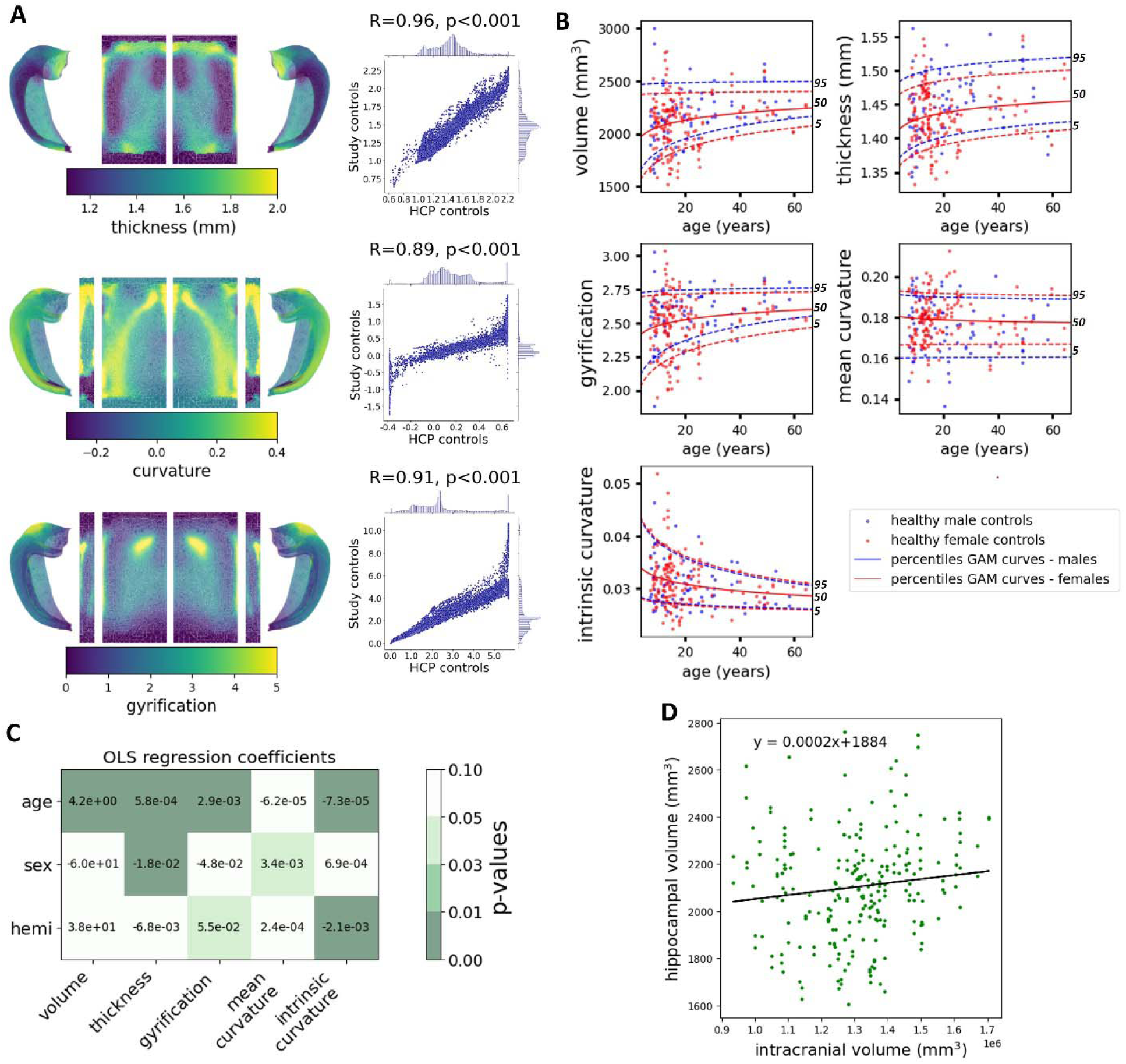
Healthy hippocampal anatomy. (A) Folded and flat maps of HippUnfold-derived surface-based thickness, curvature and gyrification in our study’s healthy controls. Correlation of surface-based features extracted from our study’s healthy controls compared to the HCP cohort. (B) Normative growth charts of harmonised features in healthy male and female hippocampi for the 5^th^, 50^th^ and 95^th^ percentiles of the population. (C) Coefficients from the linear regression model testing the effect of hemisphere, sex and age on the harmonised features and coloured by their significance (*p-values*). (D) Correlation between hippocampal volume (HippUnfold derived) and intracranial volume in healthy controls.

### Hippocampal anatomy in HS

Hippocampal volume and morphological features derived from ipsilateral hippocampi in HS patients deviated from normative curves in controls (Figure 3A). The hippocampal volume, thickness and gyrification of pathological hippocampi fell below the 5^th^ percentile range of the healthy population for 90.1%, 63.2% and 88.2% of the HS patients, respectively. Additionally, curvature and intrinsic curvature values exceeded the 95^th^ percentile in 67.1% and 70.4% of patients, respectively. Overall, in over 60% of patients with HS, all features derived from the pathological hippocampus fell outside the 5^th^ or 95^th^ percentile of the healthy population. In contrast, the distribution of contralateral hippocampi in HS and hippocampi from disease controls aligned with the percentiles of the healthy population.

**Figure 3:**
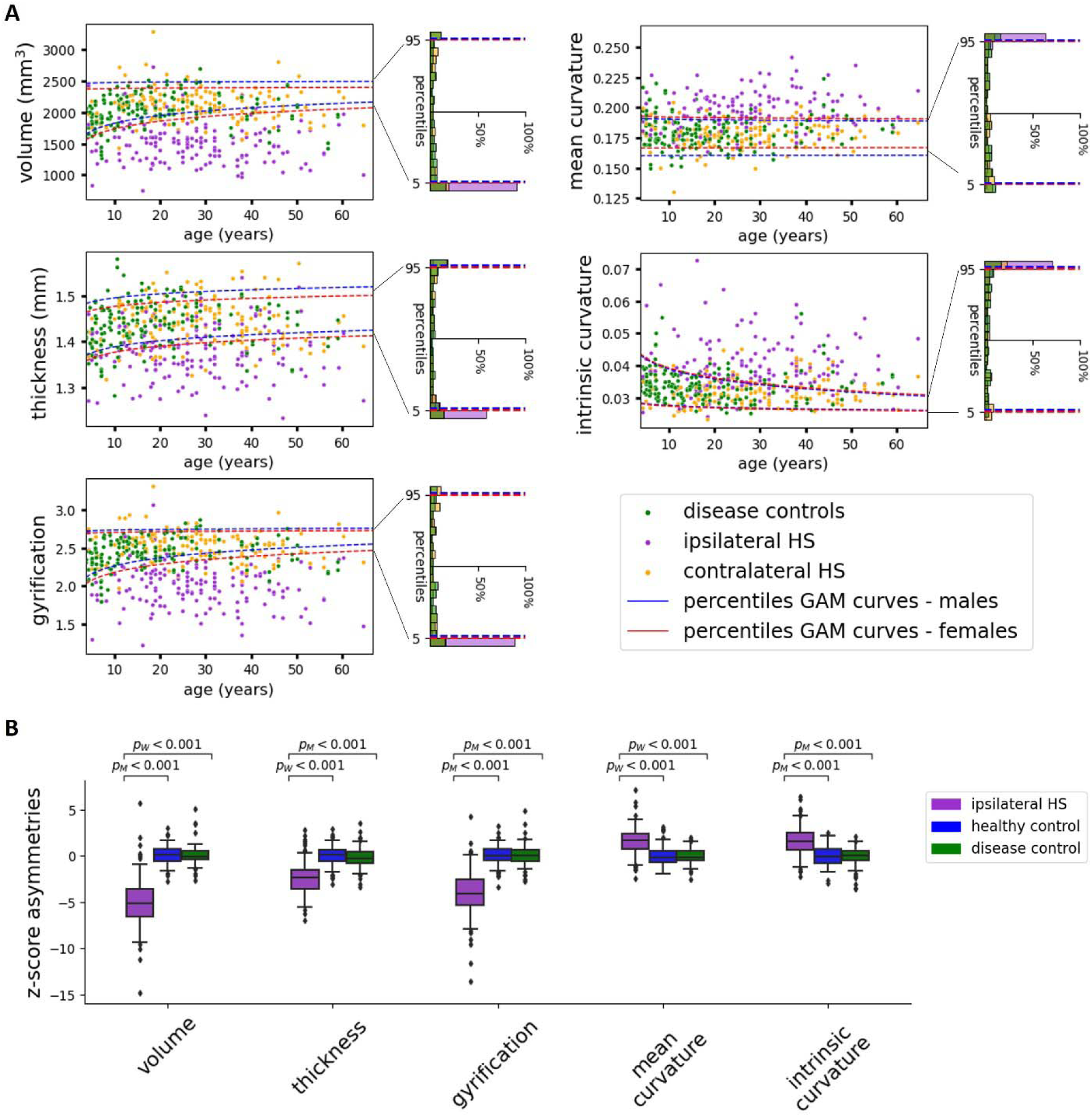
Characterisation of morphological abnormalities in HS. (A) Distribution of harmonised features in the ipsilateral hippocampi of HS patients (purple), contralateral hippocampi of HS patients (orange), and hippocampi of disease controls (green) plotted against normative trajectories derived from the 5^th^ and 95^th^ percentiles of the healthy male (blue dashed line) and female (red dashed line) controls’ hippocampi. Histograms of the percentage of each group falling within each centile of normative curves are reported on the right axis. Features from ipsilateral HS consistently fell outside of the 5^th^/95^th^ centiles for all features. (B) Boxplots of z-score asymmetries in ipsilateral hippocampi of HS patients compared to healthy controls and disease controls. Statistically significant differences in distributions between each group were assessed using the Welch T-test (p^W^) for normal distributions and the Mann-Whitney test (p^M^) for non-normal ones.

In the analysis of asymmetries (Figure 3B), patients exhibited significantly more extreme asymmetry values (all *p-values* < 0.001) between their hippocampi in all features compared to both healthy and disease controls. Ipsilateral hippocampi had reduced volume, thickness and gyrification, and increased curvature and intrinsic curvature in comparison to contralateral hippocampi. No significant differences in asymmetry features were observed between the healthy control and disease control groups.

### Automated detection and lateralisation of HS

The AID-HS classifier correctly identified 90.1% of HS patients and 94.3% of disease and healthy controls (Table 2). For HS lateralisation, the classifier accurately lateralised 97.4% of HS patients. In a subset of 24 MRI-negative patients, the classifier accurately detected 19 patients as being pathological (79.2%) and correctly lateralised 22 patients (91.7%).

Classifier performances remained consistent across the different sites and were not significantly different between age groups, with accurate detection in 91.5% of adults and 85.3% of children (*p* = 0.68) and accurate lateralisation in 98.3% of adults and 94.1% of children (*p* = 0.94). Moreover, classifier performances were not significantly different between males and females or the HS histopathological subtypes (all *p-values* > 0.05). Among controls, the classifier did not show significant differences in detecting disease controls and healthy controls (*p* = 0.27).

Overall, the AID-HS classifier trained on HippUnfold-derived features achieved 92.6% accuracy in detecting HS and 97.4% accuracy in lateralising HS. In comparison, classifiers trained solely on volumes extracted from FastSurfer achieved 88.4%-90.6% accuracy in HS detection and 95.4%-94.7% in HS lateralisation when using raw volumes and incorporating pre-processing techniques (Table 3). In summary, utilising HippUnfold-derived features with preprocessing steps, enabled accurate detection of an additional 15 HS patients and controls and accurate lateralisation in four more HS patients compared to utilising FastSurfer-derived features with and without preprocessing steps (Table 3).

**Table 3:**
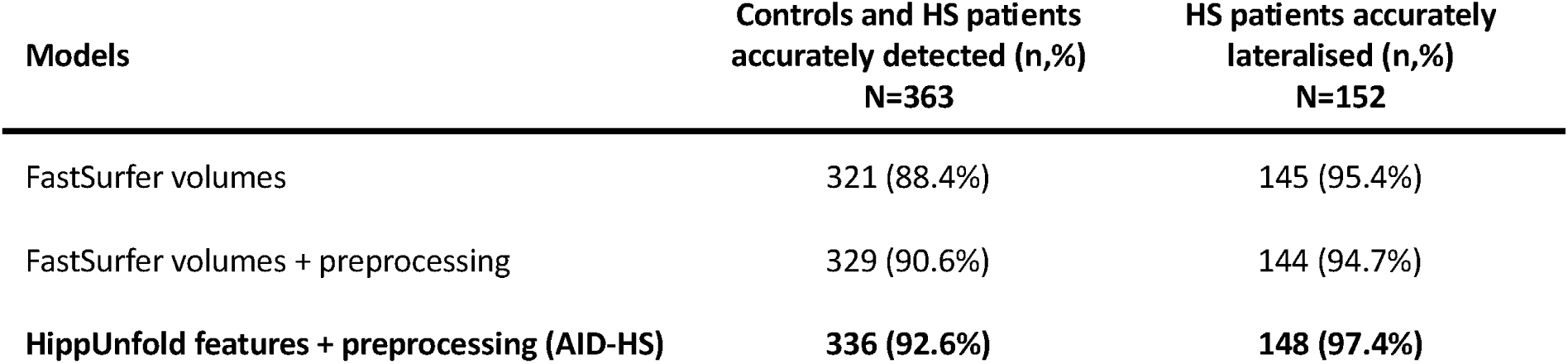
Comparison of detection and lateralisation performances across models. The best model is highlighted in bold.

### Individual hippocampal reports

In each patient, we used hippocampal feature values compared to normative growth charts, hippocampal feature asymmetry scores and results from the AID-HS classifier to create individual patient reports. Figure 4 illustrates two example reports for patients who were initially reported as MRI-negative but who were later confirmed as right (Example 1) and left (Example 2) HS through histopathological analysis. HS lateralisation in these patients was performed through intracranial EEG prior to surgery. Our reports provide a comprehensive and transparent assessment of the segmentation quality, the hippocampal morphology, and the HS detection and lateralisation scores.

**Figure 4:**
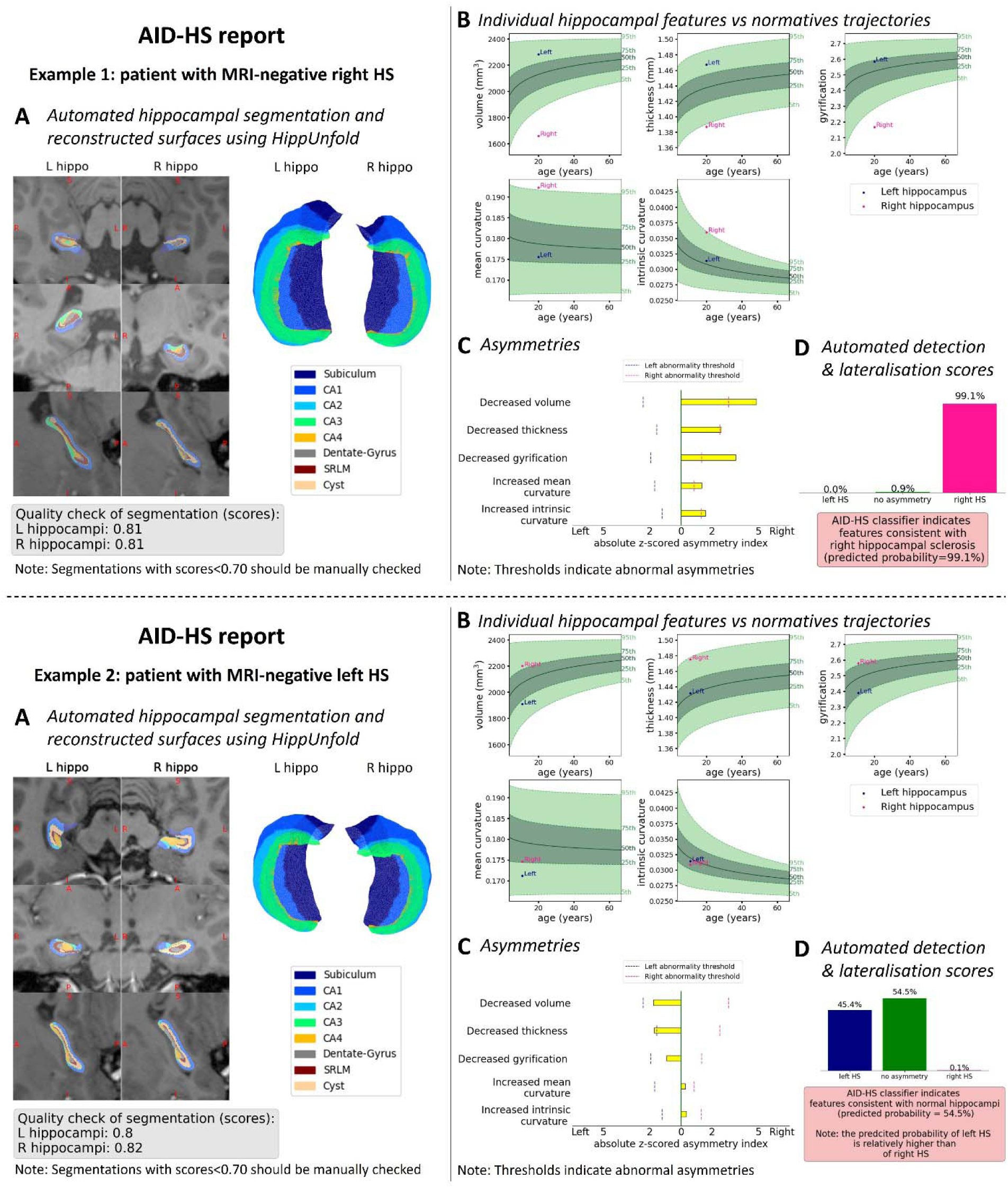
Examples of AID-HS reports for two patients with MRI-negative right HS (example 1) and left HS (example 2). (A) Automated hippocampal segmentation and reconstructed hippocampal surfaces using HippUnfold, alongside automated quality control of the segmentation. (B) Individual hippocampal features compared to normative trajectories (with 25^th−^ 75^th^ percentiles in dark green, 5^th−^ 25^th^ and 75^th−^ 95^th^ percentiles in light green, patient’s left hippocampus in blue and patients’ right hippocampus in pink). (C) Asymmetry scores against left and right abnormality thresholds (D) Automated detection and lateralisation scores from the AID-HS classifier, indicating the probability that hippocampal feature asymmetries are consistent with left or right HS or no HS.

In Example 1, automated quality control scores of 0.81 (dice overlap with template hippocampus) for both left and right hippocampi, indicated good quality hippocampal segmentations (Panel A). Compared with the normative growth charts (Panel B), the left hippocampus features fell within the normal range of the healthy population, while the right hippocampus had features that fell outside the 5^th^ and 95^th^ percentiles. In the asymmetry analysis (Panel C), abnormalities were lateralised to the right hippocampus, which had significant reductions in volume, thickness and gyrification, alongside increased curvature and intrinsic curvature. These findings were further supported by the automated classifier results (Panel D), which indicated right hippocampal sclerosis with a predicted probability of 99.1%.

In Example 2, both hippocampi had features within the 5^th^ and 95^th^ percentiles of the normal population. The analysis of the asymmetries demonstrated mixed results, with decreased volume, thickness and gyrification of the left hippocampus consistent with left HS, but increased mean and intrinsic curvature in right hippocampus, consistent with right HS. The AID-HS classifier classified this patient as having no overall asymmetry (predicted probability of 54.5%), but a higher probability for left HS (45.4%) compared to right HS (0.1%). This report is an example of a more complex patient, with normal hippocampi for the age and head size, and classified as having non-lateralising asymmetries. Nevertheless, the in-depth characterisation showed subtle atrophic, thickness and gyrification asymmetries that were consistent with left HS, which was supported by the higher lateralisation score for left HS. This illustrates how the interpretable reports could help inform clinical decision making in difficult-to-diagnose patients.

## Discussion

AID-HS is an automated and interpretable pipeline for the detection and lateralisation of hippocampal sclerosis (HS) from T1w MRI scans. We leveraged the open-source software HippUnfold to extract surface-based features and volumes of the hippocampus in a large multi-centre cohort of adults and children. We characterised the dynamic development of hippocampal morphology across a wide age range in healthy controls (seven to 60 years old). Our analysis of morphological asymmetry in HS patients revealed significant differences in the pathological hippocampi – characterised by reduced volume, thickness and density of gyrification alongside increased mean and intrinsic curvatures – compared to both healthy controls and patients with focal cortical dysplasia (FCD). These distinctive features were used for automated detection (accuracy: 90.1%) and lateralisation (accuracy: 97.4%) of HS. Notably, amongst patients with MRI-negative scans, AID-HS successfully detected and lateralised a significant proportion of HS cases (accuracies: 79.2% and 91.7%, respectively). AID-HS generates individualised reports that characterise hippocampal abnormalities and provide predictive scores for automated detection and lateralisation of HS. AID-HS is released as an open-source tool for the epilepsy community.

We extend beyond past studies in several key respects. First, our machine-learning classifier has been trained on a large, multi-centre cohort of both paediatric and adult data, with a variety of HS pathologies (HS type-1, HS type-2, HS type-3 and non-specified HS). Previous studies have often been limited to a small number of subjects from a single centre^14–16^, and trained solely using adult data^17,18^, which might limit their ability to generalise to new, previously unseen, cohorts. By training and evaluating our classifier on a heterogeneous cohort, we were able to demonstrate consistent performances across a range of ages, histopathology subtypes, and MRI scanners. In particular, by using leave-one-site-out cross-validation, we validated the ability of the classifier to generalise to new epilepsy centres, a fundamental requirement for widespread model adoption. Furthermore, previous classifiers have often relied on the use of multiple MRI modalities, such as combining T1w scans with Diffusion Tensor Imaging (DTI), T2 or FLAIR, which are not always available. The AID-HS classifier achieved state-of-the-art performance using features solely extracted from T1w MRI scans. By developing a classifier using heterogenous data from the most commonly acquired MRI protocol, we maximise the utility of AID-HS for epilepsy centres around the world.

Second, AID-HS classifier’s performance exceeded that of previous HS classifiers trained on large multi-centre data, including on MRI-negative cases. Indeed, our classifier achieved higher scores at detecting and lateralising HS (90.1% and 97.4% respectively) than the classifier trained on the large ENIGMA cohort of MRI-positive HS (75% and 83%)^17^ or a previous in-house tool for surface-based features (lateralisation only, 93%) ^18^). On a subgroup of patients with MRI-negative HS, our classifier was also able to correctly lateralise more than 90% of the abnormalities, compared to 84%^18^.

Third, AID-HS has been shown to differentiate patients with HS from patients with FCD, the leading cause of lesional MRI negative epilepsy. This is a critical capability for the presurgical planning of patients with suspected focal epilepsy with ostensibly normal MRI scans. By enabling the automated detection of even the most subtle cases of HS, AID-HS has the potential to improve post-surgical seizure-freedom rates alongside reducing the delays, burdens and costs of additional invasive investigations in patients with subtle HS.

Fourth, AID-HS was designed to create interpretable and individualised reports for clinical evaluation. While previous studies have presented promising methods for detecting and lateralising HS, none of them have offered a reusable tool specifically tailored for clinical evaluation. By openly sharing our code, packaging it into a user-friendly pipeline and providing a user-guide, we aim to support reproducibility and independent validation of our pipeline. The individualised reports have been tailored with the help of expert radiologists to fit clinical need. The main objective was to demystify the outcomes of the often-perceived “black-box” of machine-learning. Consequently, our reports deliver transparent and interpretable results, facilitating a better understanding of individual abnormalities and offering valuable information to aid the diagnosis of challenging HS cases.

Finally, this study validated for the first time the use of the open-source tool HippUnfold in 3T MRI scans acquired with clinical protocols. Indeed, we demonstrated the potential of HippUnfold to characterise both healthy development and pathological morphological changes throughout the lifespan. This validation of HippUnfold opens avenues for possible wider utility to answer other research questions, such as the characterisation of longitudinal structural change in HS, or to study other neurological diseases, such as developmental amnesia or Alzheimer’s disease.

### Limitations and future work

The HS cohort used in this study lacked examples of confirmed bilateral hippocampal abnormalities. As such we were unable to train our classifier to detect these pathologies. Nevertheless, bilateral abnormalities might be evident when hippocampi are compared to normative trajectories, and future work targeting these cases could enable the extension of these tools to bilateral HS classification.

## Conclusion

Our study introduces AID-HS, an open-source software for characterising individual hippocampal morphological abnormalities and automating the detection and lateralisation of HS in patients with temporal lobe epilepsy. By utilising features extracted from commonly acquired T1w scans in a large and heterogenous cohort of paediatric and adult patients from multiple epilepsy centres, we have demonstrated generalisable performance across a variety of individuals and sites. To facilitate independent evaluation of our software, we have made all the code available on GitHub (https://github.com/mathrip/AID-HS).

## Data Availability

All code is available online to download on GitHub

https://github.com/mathrip/AID-HS

## Supplementary material

No supplementary material

